# Time-resolving the COVID-19 outbreak using frequency domain analysis

**DOI:** 10.1101/2020.05.07.20094078

**Authors:** Keno L. Krewer, Mischa Bonn

## Abstract

Difficulties assessing and predicting the current outbreak of the severe acute respiratory syndrome coronavirus 2 can be traced, in part, to the limitations of a static description of a dynamic system. Fourier transforming the time-domain data of infections and fatalities into the frequency domain makes the dynamics easily accessible. Defining a quantity like the “case fatality” as a spectral density allows a more sensible comparison between different countries and demographics during an ongoing outbreak. Such a case fatality informs not only how many of the confirmed cases end up as fatalities, but also when. For COVID-19, knowing this time and using the entire case fatality spectrum allows determining that an outbreak had entered a steady-state (most likely its end) about 14 days before this is obvious from time-domain data. The lag between confirmations and deaths also helps to estimate the effectiveness of contact management: The larger the lag, the less time the average confirmed person had to infect people before quarantine.

## Motive

The severe acute respiratory syndrome coronavirus 2[1] is currently spreading around the world in an epidemic wave. To fight the epidemic itself as well as to mitigate collateral damage done by mass quarantine measures[2], it is key to assess the situation quickly and accurately. Much information can be already determined during the outbreak, thereby allowing to assess effectivity and necessity of countermeasures. We show here how to gain critical information using only the time series of reported confirmed and fatal cases.

## Intro

In an ongoing outbreak, the number of infected people cannot be accurately known. Mainly two numbers are communicated to describe the currently ongoing COVID-19 outbreak: the number of “confirmed cases” and the number of “deaths” [3]–[5]. The problem is that in a time-dependent situation, each number may change rapidly. It is usually more important to know the timing at which the numbers are reported than the numbers themselves.

A prime example of this is the case fatality ratio. Different timing underlies most of “The many estimates of the COVID-19 case fatality rate”[6]. In time-dependent situations, “rate” is reserved to describe quantities per unit time, so the case fatality is a ratio, not a rate.

The fraction of infected persons who end up dying should remain constant, unless radical improvements in treatment happen. This is the infection fatality ratio, an important quantity. If one has a good estimate of the infection fatality ratio in one country, one can estimate the infections in another country from the number of deaths recorded there. However, in an ongoing outbreak, the infection fatality ratio is fundamentally different from dividing the number of deaths that have occurred up to now by the number of infections up to now, because people do not die instantly from the infection. To illustrate this point, we do the following thought experiment: “A condition is fatal in 100 % of the cases. 100 people have acquired the condition by now. How many of them are dead?” The answer is: Between 0 and 100, depending on when the condition was acquired and how fast it leads to death. The timing is more important than the fatality of the condition.

In an ongoing outbreak, we cannot know the actual rate of infections; we can only know the numbers of reported confirmed cases and deaths. To understand a number, one has to understand the question it answers[7]. The number of confirmed cases reported today answers the question: “In how many cases have people been tested, confirmed positive in a laboratory, with this confirmation having been reported today?” This question is quite complicated. The number of confirmed cases depends on several factors. The actual number of infected people is only one of them; usually the number of tests[8] and the day of the week are important. The number of deaths answers the question: “How many of the confirmed infected appear to have died from COVID-19?” This is a bit simpler, but the “appear” does leave room for interpretation. The problem with finding the infected is that many present very mild symptoms that are indistinguishable from those associated with influenza and other common respiratory diseases usually summed up as “the common cold”. The severe cases, especially those leading to deaths, are harder to miss. Therefore, Ward[8], and Flaxman et. al.[9] conclude that the reported deaths are likely closer to the actual deaths than the confirmed are to the actual infected. If one wants to fight a pandemic, not merely monitor it, another question becomes important: “In how many cases do we know that infectious people stopped spreading the disease because they were put into quarantine?” That number, too, is the number of confirmed cases, since confirmed infectious persons generally will be quarantined.

## So

How can we describe the time dependence of the observables of an epidemic?

The problem is that the confirmations *C* reported on the day *t* depend on all infections *I* on each day before that day *t* and how long before they happened. Mathematically:

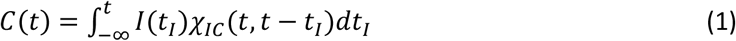

Here *X_IC_*(*t,t − t_I_, x*) is the infection confirmation response function, which gives the probability at time *t* of reporting the confirmation of an infection that happened *t* − *t_I_*, days before *t_I_* is the time of infection. In the following, we will assume that this probability does not change over time, and hence only depends on how much earlier the infections happened:

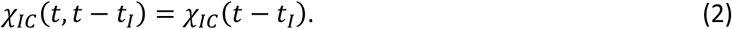

The same can be done for the death rate *D*(*t*) at time responding to the confirmation rate *C*(*t_C_*) at time *t_C_*:

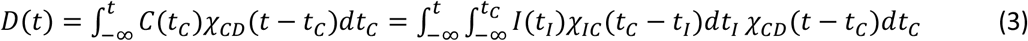

*χ_CD_*(*t − t_C_*) is the case fatality response function. We note that even under the simplifying assumptions of no changes in testing, reporting, or treatment of the disease over time, we are left with complicated convolution integrals over the infections, which themselves change exponentially over time. About 200 years ago, Fourier[10] faced the same problem in the description of heat transport, where heat and temperature also change exponentially over time (and space). His work lead to the development of the Fourier transformation, which simplifies the description of a time-dependent phenomenon *B*(*t*) by transforming it into a spectral density 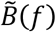 in the frequency (*f*) domain:

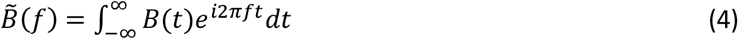

Fourier transforming equation (2) replaces the convolution in the time domain with a simple multiplication in the frequency domain.

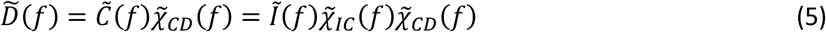

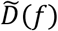, 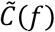 and *Ĩ*(*f*) are the deaths, confirmations, and infections happening at a frequency 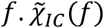, 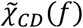 and 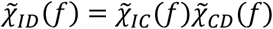 are the infection confirmation ratio, the case fatality and the infection fatality, respectively. So one can simply divide fatal cases by confirmed cases to obtain the case fatality ratio – in the frequency domain. This simplicity comes at a price, though: The case fatality is not a single number, but an entire spectrum of multiple frequencies. Further the case fatality 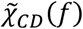 at each frequency *f* is a complex number^1^ composed of an amplitude 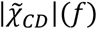 and a lag *τ*(*f*):

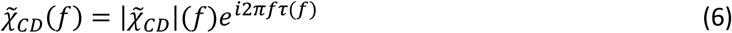

The amplitude of the case fatality 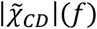 answers the question: “What fraction of the confirmed cases reported at frequency *f* end up dying?” And the lag *τ*(*f*): “How soon?”

Let us apply this formalism to reported data. For the initial demonstration, we chose a place where the outbreak is over and the reporting and testing policy did not substantially change over time: China, excluding Hubei province.

We take the daily reports of confirmations and deaths from the data published by John Hopkins University on GitHub[3], [11]. Dong, Du and Gardner’s[3] data start on January 22^nd^, 2020, which is day t=0 in this paper. One can already see in the time domain data in fig. 1 a) that the deaths trail the confirmations by about 10 days. Fourier’s formalism implies continuous observation over time, while we here have discrete observations, one each day. A discrete Fourier transformation algorithm now called fast Fourier transform was invented by Fourier’s contemporary Gauss[12], [13] to analyse the time-dependent observations of planets and comets. The fast Fourier transform has become extremely common; last but not least digital copies of this paper may be compressed using it. The compression works because the most relevant information is contained in very few frequency steps. In the case of China ex. Hubei, the amplitude spectra of confirmations and deaths in fig. 1 b) start fluctuation randomly for frequencies larger than 0.06/day. This means data at these frequencies are most likely dominated by statistical fluctuations and do not contain much useful information. We can therefore limit our analysis to the 6 frequency steps below 0.06/day and still capture the relevant information from two series of almost 100 time steps (days). Hence, we only plot the case fatality for these first frequency steps in fig. 1 c). We can see that the amplitude and the lag are quite constant, at almost 0.9 · 10^−2^ fatalities/confirmation and ca. 11 days, respectively. This means the outbreak in China ex. Hubei can be described in the simple terms that about 1/100 of confirmed cases died, and the death was reported on average 11 days after the confirmation. This, however, could also have been inferred by just overlapping the curves of confirmations and deaths, albeit less mathematically rigorous. We aim to retrieve a good estimate of the case fatality in an ongoing outbreak, not just in one that is essentially over. We stick with China ex. Hubei and ask the question: “What could we have known on day 20 (February 11^th^)?” Well, when we Fourier transform the rates reported up to day 20, we see in fig. 1 c) that the 0 frequency value of the case fatality is 0.35 · 10^−2^ fatalities/confirmation, much lower than the final value. This is not surprising, because the 0-frequency value is just the accumulated number of deaths divided by the accumulated number of confirmations. As explained above, this value is quite useless as the outbreak is ongoing on day 20.

**Fig. 1.**
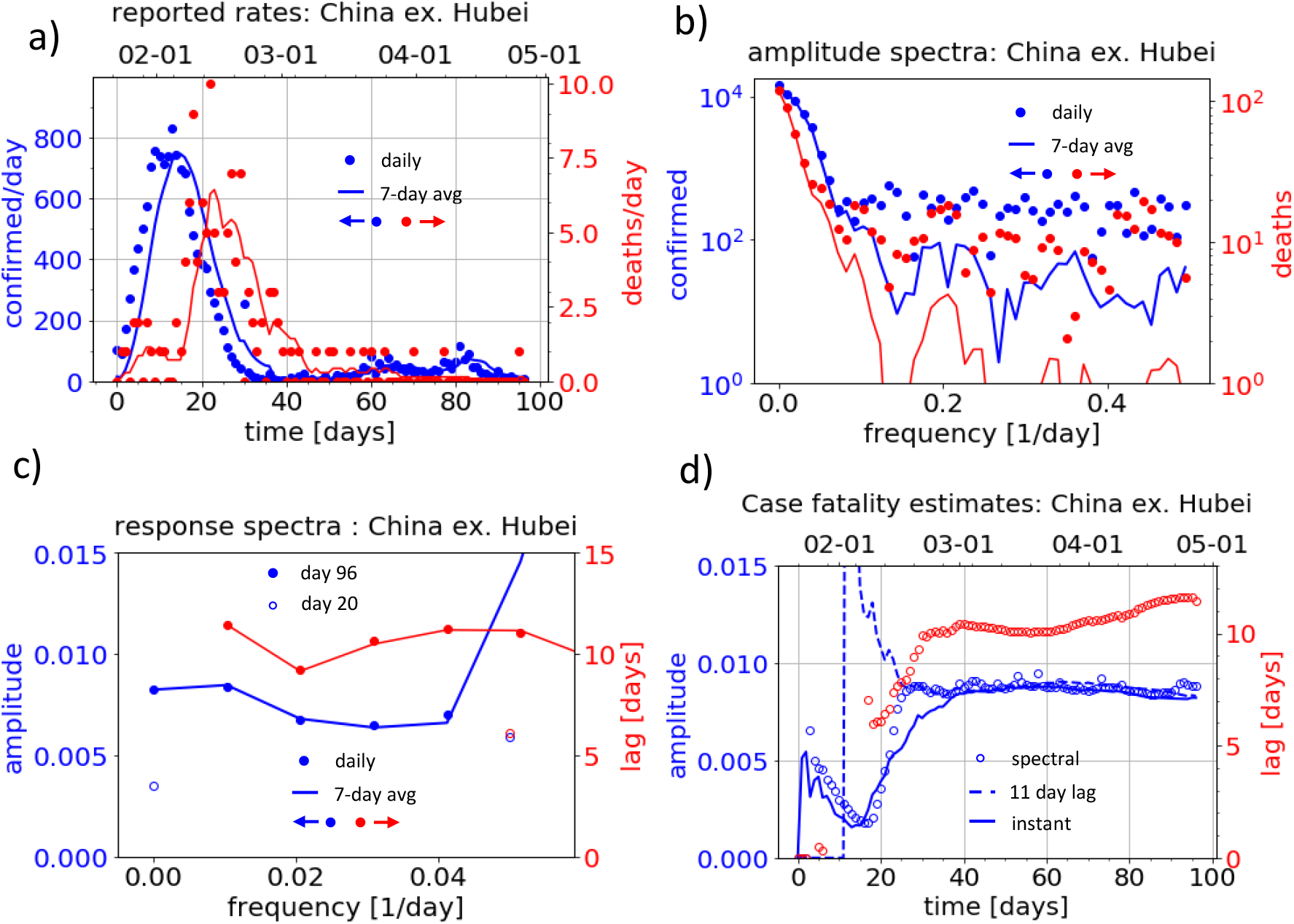
a) Reported rates of confirmations (blue, left axis) and deaths (red, right axis) in mainland China, excluding Hubei. Dots are daily reports, lines are floating averages over the past 7 days. b) Amplitude spectra of the time traces from a). c) Case fatality amplitude (blue, left axis) and lag (red, right axis) for the frequencies above the noise floor, which is below 0.06/day. Full dots come from dividing the spectra from day 96, seen in b); empty dots from spectra derived from the first 20 days. d) Estimates of amplitude and lag of the case fatality by different methods as a function of time: Full lines are deaths reported up that time divided by confirmations up to the same time. Using the confirmations obtained 11 days earlier yields the dashed line. The dots are obtained from spectra taken up to that time, the amplitude is a weighted spectral average of based on 7-day averaged spectra, the lag is the value at first frequency above 0 from daily spectra.

The second Fourier component, however, is already at 0.6 · 10^−2^ fatalities/confirmation, much closer to the final value. In general, we should average over the whole spectrum. We suppress the noise from statistical fluctuations by first using a 7-day floating average over the daily reports and then weighting the average by the spectral intensity of deaths, since the lower number of deaths, the larger their relative statistical error. The 7-day floating average unfortunately delays the time traces by half a week, costing some valuable time and reducing time resolution. It is necessary for estimating amplitudes, especially in countries which report significantly less on weekends. Even with the delay from the 7-day average, the spectral average converges towards to final case fatality value around day 25, about two weeks before the “static case fatality” does.

We also have *a posteriori* recognized that deaths lag the confirmations by about 11 days. When we compare the deaths accumulated up to a certain day with the confirmations accumulated up to 11 days earlier, we also get an estimate of the case fatality amplitude that converges towards the final value after day 25. While we only know this 11-day value a posteriori from our Fourier analyses, one could infer it from studying individual cases much earlier: Wang et al.[14] reported a median time between the onset of first symptoms and the onset of the acute respiratory distress syndrome of 8days on February 7^th^ (day 16). The first symptoms set in shortly after a patient can be tested positive and respiratory distress is how most severe acute respiratory syndrome corona virus 2 patients die, at least those dying quickly. So those 8 days would have given reasonable initial guess for the lag.

Having obtained the case fatality of the outbreak in China ex. Hubei, we can now answer the question: “How many confirmations *C_X_*(*Y*) would an ex. Hubei style system have reported at if it reported deaths like country Y?” This is done by dividing the spectrum of deaths 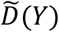 of country Y by the case fatality 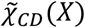 of China ex. Hubei. (X here stands for China ex. Hubei).

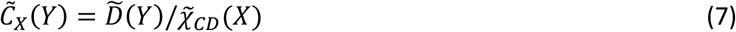

This gives the confirmation spectrum 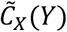 for county Y assuming the case fatality of region X. An inverse Fourier transform then yields the confirmation rates *C_X_*(*Y*). We use 7-day floating averages to suppress statistical noise. The most important assumption for the merit of this comparison is that deaths and confirmations behave linearly with respect to each other, i.e., that more confirmations do not lead to a change in case fatality. We call this comparison “ex. Hubei standard” and perform it for two places which perform widespread testing and where the outbreaks are more recent: South Korea (seen in fig. 2) and Germany (fig. 3). In summary, the “ex. Hubei standard” calculates the number of infected from the number of deceased, using the case fatality of China.

**Fig. 2.**
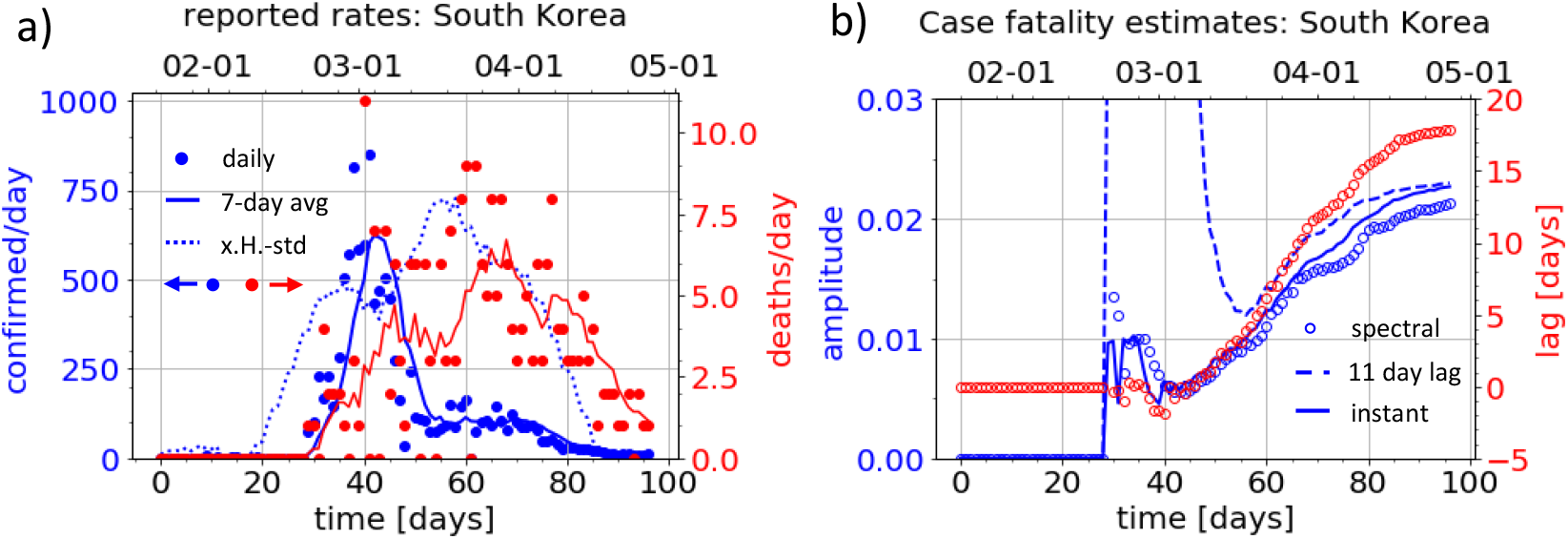
a) Reported rates of confirmations (blue, left axis) and deaths (red, right axis) in South Korea. Dots are daily reports, full lines averages over the past 7 days, and the dotted line are confirmations in an ex. Hubei type response based on the 7-day averaged deaths in South Korea. b) Estimated case fatality amplitude (blue, left axis) and lag (red, right axis) based on data

**Fig. 3.**
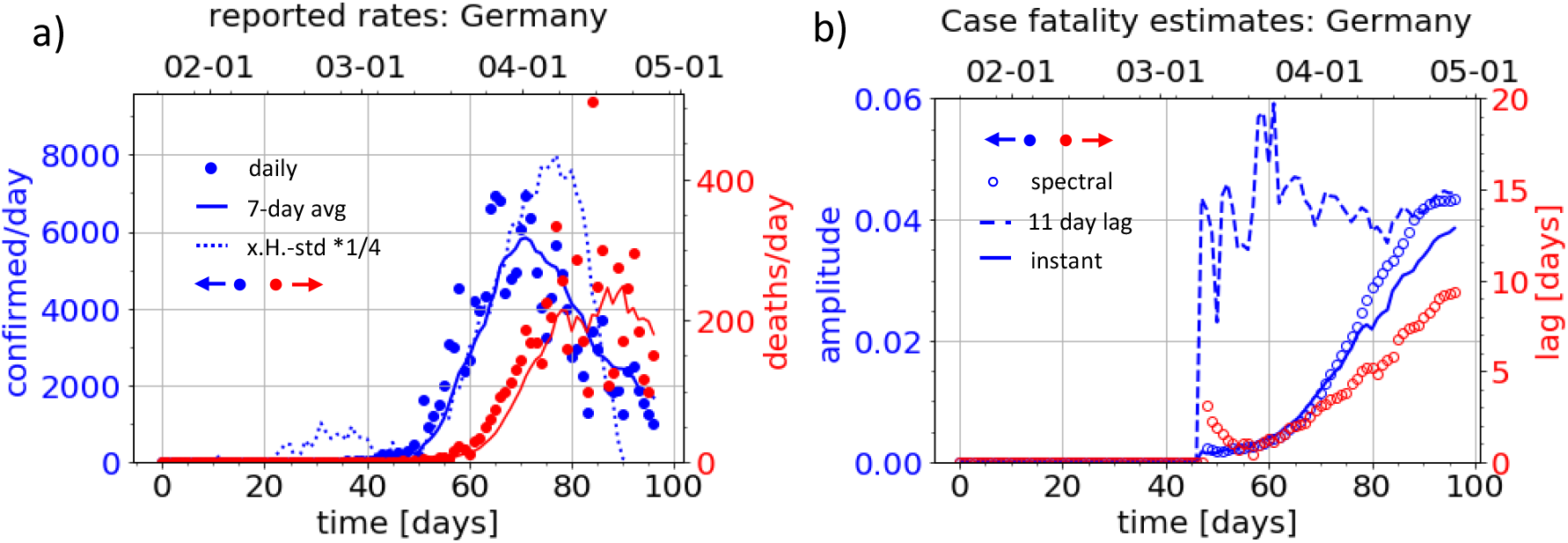
a) Time traces of reported rates of confirmed (blue, left axis) and deaths (red, right axis) in Germany. Dots are daily reports, full lines averages over the past 7 days, and the dotted line are confirmations in the ex. Hubei standard divided by 4 for scale. b) Case fatality amplitudes (blue, left axis) and lags (red, right axis) estimated using data available at the respective time. The full line denotes the instantaneous ratio of accumulated fatalities and confirmations, the dashed line uses confirmations accumulated until 11 days prior, and the dots result from spectral analyses.

For Korea, the ex. Hubei standard shows an increase to a hundred possible confirmations per day around day 25, ca. 10 days before the Koreans actually find and confirm several hundreds of infected each day. When the Koreans do, though, they find more than the Chinese would have in the same time period. This indicates that a few thousand infected people had gone unnoticed for ca. one week, but then the Koreans identified most of them. This fits the magnitude and timeline of the Shincheonji cluster[15]. After this initial trend, the ex. Hubei standard based on the deaths in Korea again indicates much more possible confirmations than the Koreans report.

How can we understand these discrepancies? There can be two reasons: a) The case fatality is nonlinear, which invalidates eq. (2). b) The case fatalities between Korea and China are fundamentally different. Discrepancies from day 25 to day 45 can be explained by non-linearity: The Korean’s employed rapid contact tracing, which means a single confirmation at a time triggers several confirmed contacts soon after. This allows confirmations (and, more importantly, quarantine of infected) to outpace infections, beating exponential growth by faster exponential growth. We note that contact tracing only leads to nonlinearity of the overall response if no new hidden clusters continuously form to be contact-traced later. Nonlinearity is not a good explanation for the discrepancies after day 45, as no surges in confirmations happened after the ex. Hubei standard alleged further possible confirmations in Korea. This would imply Korea having lost most of its capabilities to confirm cases, while still managing to curb the spread of the disease. We consider this unlikely and therefore we search for possible causes for differences in the case fatality. We look at how the case fatality of South Korea differs from China ex. Hubei. In South Korea about twice as many people (2.2 · 10^−2^ compared to 0.9 · 10^−2^) people die after confirmation, but they also die much later than in China. This contradicts what we would expect from either more infections among the risk group or worse health care; in either case more people should die <u>sooner</u> but we observe that more die <u>later</u>. Similarly, false positive confirmations in China cannot explain the observed combination amplitude and lag. A complicated interplay between those factors cannot be ruled out as an explanation without very detailed data. A simpler and therefore better[16] explanation for the observed discrepancy is that China reports only the cases that have certainly died from the severe acute respiratory syndrome coronavirus 2, while South Korea reports everyone who died while not having yet recovered from the virus. A rapid disease progression is characteristic for the severe acute corona virus [14], [17], hence one would expect characteristic cases to die quickly, as they do in China. The risk groups for a severe case of coronavirus infection are elderly people with pre-existing health conditions[17]. These are people with a low remaining life expectancy. This means many risk group patients would be expected to die from other causes than the coronavirus before they would have had time to fully recover. It takes three to six weeks for severe cases to recover [18]. For a group of random people with an average life expectancy of 2 years (without CoViD-19 infection) we can estimate that a fraction of 4 · 10^−2^ of them will die within one month^2^. This makes differentiating between a death from Corona virus and “random” death increasingly difficult, the later the death occurs. China only reporting the quick deaths while Korea also reporting slow deaths, which may not have been caused by the infection, is the simplest explanation we can find for the discrepancy.

We now turn to Germany and compare the numbers with the cases in China ex. Hubei. We can see that the curve of confirmations in Germany has a very similar shape as the ex. Hubei standard would predict, but the number of confirmations is ca. 4 times lower than in China. The Germans found about 1/4 of the cases that the Chinese would have found, provided they reported the same amount of deaths. Since the German timing seems to be very similar to the Chinese (ex. Hubei) it is not surprising that the case fatality amplitude corrected by the lag from China ex. Hubei has fluctuated around a constant of 4.4 · 10^−2^ since the first death in Germany. By now the spectral averaged case fatality has reached a similar level. This example illustrates the futility of using the instantaneous case fatality ratios as the case fatality amplitude of about 4 · 10^−2^ in Germany at day 50 was expected when accounting for the lag known from China, while the instantaneous case fatality lay at 0.1 · 10^−2^. We note that a day change in the lag would have resulted in an absolute change in estimated case fatality amplitude by 1 · 10^−2^ at day 50, but now it will only change by 0.1 · 10^−2^ for a 1 day different lag. We note that at this point, most of the data is still from the rising flank of the outbreak, hence we cannot distinguish if Germany is reporting patients dying very late similarly to Korea or to China, since most late deaths have not occurred yet. While a sizable fraction of deaths has yet to occur in Germany, very little new infections will (provided no major changes are induced in the behaviour of the Germans). This is what the observations of constant spectral case fatality estimates in recent days tell us. Germany is entering a steady-state, as did China on day 25. Since we have observed that the outbreak is essentially over, provided no major change is made in Germany, how can we monitor if a major change happens, i.e. if the outbreak is restarted by lifting strict social distancing measures?

Death reports occur too late to be useful, confirmed cases depend more on the effectiveness of the testing scheme than on the number of infections[8], and as long as most of the infections are confirmed quickly, contact tracing and quarantine suffices to stop the spread of the disease. We need to tell if enough people have been found and quarantined quickly enough. Is there a single value that can be easily reported, which will tell if a new surge of COVID-19 infections is happening or if contact management is working? The answer to the ultimate question about COVID-19, the contact management and all the rest is: “- 5 days”. The exact question is: “How long was the average time between onset of symptoms and quarantine for the cases confirmed today?” We count here the ability to produce a positive test as a symptom. 5 days is a recent estimate of the average incubation time [18]. If most infected have been quarantined before they became infectious, the average time between symptoms and quarantined must be below 0 and cannot go lower than (minus) the average incubation time. We urge to focus on reporting this time, rather than the precise numbers of confirmations.

Measuring timescales is more important and reliable than quantifying the time-dependent observables in dynamic situations, since observables will change drastically over time, but timescales tend to be determined or at least limited by underlying time constants, in this case the incubation time. This is the underlying reasoning how we come up with the “ultimate question” and the answer.

Can we tell this lag between infectiousness and quarantine from our current analysis? No. But we can give an indication of where the lag between infectiousness and quarantine was smallest for the past confirmed cases: We can expect cases to be quarantined by the time they are reported, and the average time between infection and death is another time constant of the disease^3^. So, the larger the lag between reported confirmations and deaths, the smaller the lag between infections and quarantine must have been. We plot the current lag and amplitude estimates for the case fatality for all countries with more than 100 deaths from COVID-19 in fig. 4. The absolute magnitudes of the case fatality mostly tell how widely a country has been testing[8]; the more tests, the lower the amplitudes. From the case fatality, we can gauge the state of the outbreak at the time when the people dying now had been infected: when the 11-day-lag corrected case fatality (marked x) and spectral average of the amplitudes (marked ×) have become similar, the outbreak was entering a steady state; the infection rate was past its peak. This has by now happened in Italy, Belgium, Germany, Iran and China, to name the examples with the highest fatality count. The lag allows us to differentiate between 3 testing schemes: Germany and China ex. Hubei have lags on the order of 10 days and few fatalities per case, because they managed to even test many people with mild symptoms relatively soon. Italy and Belgium had restricted testing mainly to suspected cases with severe symptoms. Since severe symptoms are fewer and take ca. 4 days [14] to develop, lags are below 5 days, and case fatalities are several times larger. However, this testing policy has been somewhat consistent throughout the outbreak. Hubei and Iran are the third type of response. Here the lag is negative. Confirmed cases were only widely reported after people had already started dying. Most likely, these territories responded to deaths by increasing testing and reporting. Lags may also be negative in very early stages of the outbreak, when the 11-days-corrected estimate may massively overestimate the case fatality while the spectral average massively underestimates it. This can be seen in the early stages of the Korean timeline in fig. 2 b). The data in fig. 4 indicates that for example Ecuador, Morocco, Bangladesh and Saudi Arabia are currently in this early stage of their respective outbreaks.

**Fig. 4.**
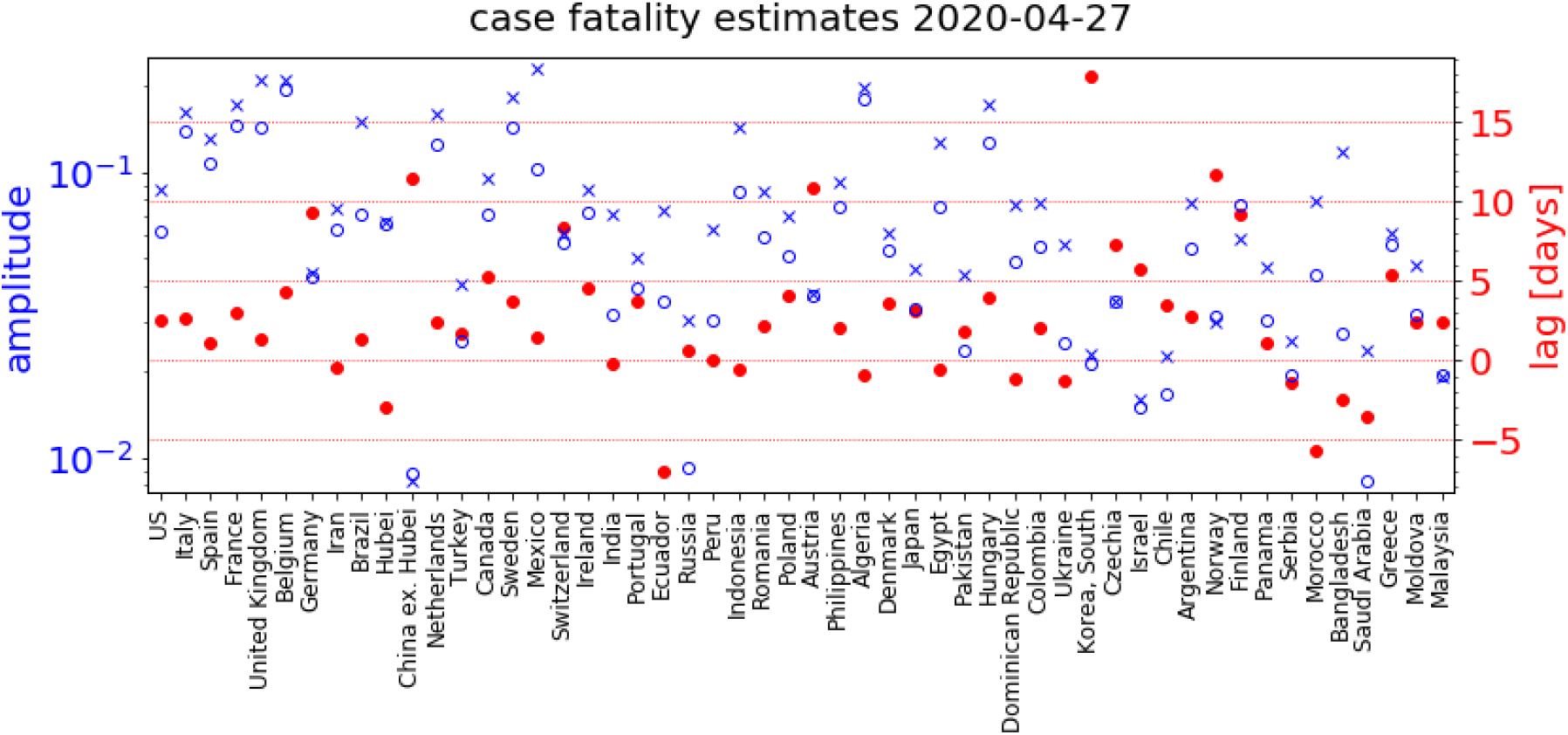
Amplitude (blue, left axis, logarithmic) and lag (red, right axis) of the case fatality for territories with more than 100 deaths. The amplitude is estimated in two ways: fatalities divided by the confirmations 11 days prior are marked with x, averages of the amplitude spectrum with o. When those two estimates start matching, the outbreak was entering a steady state and much fewer infections happened than during its beginning, the latter happened one average infection-fatality lag prior. A large case fatality lag indicates a low infection quarantine lag, which means the confirmed cases had little time to infect other people before being quarantined.

South Korea was probably the only country that got its initial outbreak under control mainly by contact management rather than social distancing. By now, however, many countries should have the test and contact management infrastructure to do the same. Countries with a lag close to 10 days were already within reach of this goal before. They can switch to this strategy now and monitor the situation by reporting their answer to the ultimate question: “How much time did the infectious people have to infect more people?” This can even be done in countries that do not have enough test resources, by quarantining all even mildly symptomatic people and their contacts on suspicion and only test a small fraction of them, preferably those without known epidemiological links to confirmed cases. It may be more important to test and report quickly and smartly rather than extensively to get a reliable and timely estimate for the average time a recent infectious case has spent unquarantined, and this time is more important than the absolute number of past infections.

## Conclusion

Analysing static quantities like the accumulated number of confirmed cases and deaths is not particularly helpful in understanding a dynamic situation. Fourier analysis of the time series of confirmation and death rates yields the case fatality spectrum, which allows a more sensible comparison between different places at different stages of their outbreaks. For example, in comparing China ex. Hubei and South Korea, we could tell the existence, timing, and magnitude of the Shincheonji cluster from the confirmation and death rates alone. We further conclude that the main difference in case fatality between South Korea and China ex. Hubei was reporting, most likely of deaths, since this is the only explanation for the discrepancies both in the fraction confirmed infected who die and the lag between confirmations and deaths. Further, we can tell when the static description converges towards the Fourier description that includes dynamics. Thereby, we know when the outbreak has been ending. This has, by now, happened in most severely affected countries. The key to understanding a dynamic situation is to know the time constants involved. Fourier analysis allows inferring some information on the average time a confirmed case had to infect more people, but we can do this only based on the number of deaths, which means the most recent infection situation we can assess that way is at least 2 weeks outdated. We recommend reporting a more up to date and useful quantity in a dynamic outbreak: the average time between infectiousness and quarantine for the recently confirmed cases. This time allows assessing the situation while at the same time indicating how recent the assessment is, and it can be as recent as the incubation time permits.

## Data Availability

We use the raw data published by the Center for System Science and Engineering of the John Hopkins University.
The python code for the data analyses is published in an Edmond repository.

https://github.com/CSSEGISandData/COVID-19/tree/master/csse_covid_19_data/csse_covid_19_time_series

https://edmond.mpdl.mpg.de/imeji/collection/VVStKlQ0xKllTTkH

## Supplementary Information

The python program used to perform this analysis and create the plots is available under: https://edmond.mpdl.mpg.de/imeji/collection/VVStKlQ0xKIITTkH

## Footnotes

1 We use the ^~^ to indicate all complex numbers in this paper.

2 Under the assumptions of random and uncorrelated deaths

3 Well, differences in treatment and especially reporting of late deaths may change it, as we discussed for Korea and China, but not by an order of magnitude.

